# Vascular and neurodegenerative contributions to cognitive decline and multidomain progression in de novo Parkinson’s disease

**DOI:** 10.64898/2026.09.09.26362673

**Authors:** Roqaie Moqadam, Houman Azizi, Shima Raeesi, Yashar Zeighami, Mahsa Dadar

**Affiliations:** Department of Medicine, University of Montreal, Montreal, Quebec, Canada; Douglas Mental Health University Institute, Montreal, Quebec, Canada; Centre de Recherche, Institut Universitaire de Gériatrie de Montréal, Québec, Canada; Montreal Neurological Institute and Hospital, McGill University, Montreal, Quebec, Canada; Integrated Program in Neuroscience, McGill University, Montreal, Quebec, Canada; Department of Psychiatry, McGill University, Montreal, Quebec, Canada

**Keywords:** Parkinson’s disease, cognitive impairment, cognitive conversion, deformation-based morphometry, white matter hyperintensities, cognitive decline, longitudinal neuroimaging

## Abstract

**INTRODUCTION:** Cognitive decline in Parkinson’s disease (PD) may reflect interacting neurodegenerative and cerebrovascular processes, but their distinct temporal contributions remain unclear.

**METHODS:** Newly diagnosed, untreated PD participants from the PPMI who were cognitively normal at baseline and had at least three years of cognitive follow-ups were included. Linear mixed-effects models examined longitudinal atrophy, white matter hyperintensity (WMH), and clinical trajectories. Mixed-effects models also examined the associations between baseline WMHs and a decline-related atrophy pattern expression score with subsequent multidomain clinical progression.

**RESULTS:** 58 participants experienced cognitive decline and 292 remained cognitively stable. Converters showed faster decline across all cognitive domains, accompanied by faster worsening of motor, gait, neuropsychiatric, sleep-related, and functional symptoms. Converters further showed higher baseline WMH burden without accelerated accumulation, alongside accelerated limbic and subcortical atrophy. Greater baseline WMH burden was associated with differential longitudinal progression in converters across eleven clinical outcomes, including cognition, gait, and functional independence (all q<0.05). Baseline atrophy pattern expression showed similar converter-dependent associations with longitudinal progression across eight outcomes, with the strongest interaction for MoCA (q<0.001). In joint models, atrophy pattern expression retained broader associations with cognitive and motor progression, whereas WMH burden retained more selective associations with neuropsychiatric symptoms and functional independence.

**DISCUSSION:** Cognitive decline in PD was characterized by accelerated neurodegeneration occurring against a background of WMH-related cerebrovascular vulnerability rather than accelerated WMH accumulation, supporting distinct temporal patterns and complementary clinical associations of neurodegenerative and vascular processes with cognitive impairment and broader disease progression in PD.

**Highlights:**

- PD converters showed accelerated limbic and subcortical gray matter atrophy.
- WMH burden was higher in future PD converters.
- Baseline atrophy pattern expression predicted cognitive and motor progression.
- WMH burden was selectively associated with neuropsychiatric symptoms and functional independence
- Vascular and neurodegenerative markers provided complementary prognostic information.

**Research in Context:** *Systematic review:* The authors reviewed peer-reviewed literature on cognitive decline in Parkinson’s disease (PD), emphasizing longitudinal structural MRI, white matter hyperintensities (WMH), and studies examining clinical progression. Prior work supports both neurodegenerative and vascular contributions to cognitive decline, but their longitudinal relationships with cognitive decline and multidomain progression remain incompletely characterized.

*Interpretation:* While both WMHs and neurodegenerative pathologies contribute to cognitive and multidomain impairments in PD, WMH burden differences precede symptomatic cognitive decline, whereas longitudinal progression of atrophy is more closely intertwined with cognitive decline.

*Future directions:* Independent longitudinal cohorts should examine the interactions between longitudinal progression in WMHs, neurodegeneration, molecular biomarkers, and clinical symptoms and whether their combination can improve individualized prediction of cognitive decline and subsequent progression to dementia in PD.

## 1. Introduction

Parkinson’s disease (PD) is a progressive neurodegenerative disorder traditionally characterized by its motor symptoms. However, cognitive impairment has been recognized as one of PD’s most common disabling non-motor features^1,2^. A significant proportion of individuals with PD experience mild cognitive impairment (MCI) and dementia throughout the course of the disease, with dementia affecting 25-30% of patients at any given time and developing in the majority of those surviving more than 10 years after diagnosis^1^. Importantly, cognitive trajectories show significant heterogeneity in PD patients^1,3^. While some patients maintain cognitive stability for years, others show progressive decline across multiple cognitive domains^1,2^. Understanding the neurobiological mechanisms that precede and accompany cognitive decline in PD remains a critical challenge for disease prognosis and subtyping, and designing targeted interventions^2–4^.

Prior neuroimaging studies have identified structural brain changes associated with cognitive decline in PD^4–7^. Cognitive impairment in PD is associated with accelerated cortical thinning and progressive regional atrophy, with distinct patterns of degeneration emerging across cognitive stages^2,4,7–10^. Longitudinal and prospective neuroimaging studies have implicated the thalamus, striatal regions, including the caudate and nucleus accumbens, limbic structures and medial temporal lobe regions^11^ in cognitive deterioration in PD^4,6,12^. Prospective studies have identified abnormalities in the amygdala and thalamus before clinically evident cognitive decline^6,11^.

White matter hyperintensities (WMHs) are MRI markers of cerebral small vessel disease. While WMHs are frequently observed in PD, their contribution to cognitive impairment remains controversial^13,14^. A systematic review and meta-analysis showed that WMHs are not globally more prevalent or severe in PD than age-matched controls, suggesting that WMHs are not a disease-specific feature of PD^13^. However, WMH burden was higher in PD dementia compared with cognitively normal PD patients, suggesting that WMHs may be more closely related to cognitive stage than to PD diagnosis per se^13,15^. In de novo PD, however, an early cross-sectional study found no significant association between WMH burden and cognitive function^16^. In contrast, longitudinal studies suggest that WMHs may modify subsequent cognitive trajectories, with greater baseline WMH burden associated with faster cognitive decline and progression to more severe cognitive impairment in PD^17–20^. Whether faster longitudinal WMH accumulation results in subsequent cognitive decline in PD remains incompletely understood^21^.

Despite this evidence, the relative contributions of pre-existing atrophy and accelerated neurodegeneration during the transition to cognitive impairment remain incompletely characterized^4,7–10,12,22,23^. Second, few studies have examined cerebrovascular and neurodegenerative abnormalities simultaneously, whether they show distinct temporal relationships with cognitive decline remains unclear^20,24^. Furthermore, cognitive impairment in PD is associated with progression across multiple clinical domains rather than isolated cognitive deterioration. Longitudinal studies have linked cognitive vulnerability to a broader progressive phenotype encompassing motor and gait deterioration as well as neuropsychiatric, autonomic, and sleep-related abnormalities^25–27^. Neuropsychiatric symptoms, including anxiety and depression, have also been associated with subsequent cognitive decline in PD^28^.

Although cerebrovascular injury and neurodegenerative structural changes may represent partially distinct sources of vulnerability^20,24^, existing prognostic studies have generally examined selected clinical outcomes or specific imaging measures^17,20,29^. As such, the extent to which baseline vascular and neurodegenerative abnormalities provide overlapping or complementary information regarding subsequent multidomain progression remains poorly characterized. Examining these processes jointly within a longitudinal framework of cognitive conversion in PD may therefore clarify both their temporal roles in cognitive conversion and their complementary contributions to broader clinical heterogeneity.

In this study, we investigated the longitudinal trajectories of structural brain changes associated with the transition from normal to impaired cognition in a cohort of newly diagnosed PD participants. Our aims were to i) identify potentially differential rates of longitudinal regional brain changes associated with cognitive decline in PD, ii) characterize corresponding differences in longitudinal trajectories across cognitive, motor, neuropsychiatric, autonomic, sleep-related, and functional domains, and iii) determine whether vascular and neurodegenerative changes provide overlapping or complementary information regarding subsequent multidomain clinical progression when examined jointly.

## 2. Methods

### 2.1 Study population

MRI and clinical data were obtained from 982 participants with PD from the Parkinson’s Progression Markers Initiative (PPMI) database (www.ppmi-info.org/data), a longitudinal, multi-site observational study of newly diagnosed and untreated patients with PD across the United States and Europe^30^. Only PD participants who were cognitively normal at baseline and had a minimum of 3 years of longitudinal cognitive follow-ups available were included. Participants were excluded if they were not de novo (i.e. were receiving dopaminergic medication) at baseline, or had missing MRI, demographic, or clinical data.

### 2.2 Clinical assessments

Demographic and clinical variables of interest included age, sex, years of education, global cognitive performance (Montreal Cognitive Assessment [MoCA])^31^, motor severity (Movement Disorder Society Unified Parkinson’s Disease Rating Scale Part III [MDS-UPDRS III])^32^, tremor, postural instability and gait difficulty (PIGD)^33^, Hoehn and Yahr disease stage^34^, apathy^32^, depressive symptoms (15-item Geriatric Depression Scale [GDS-15])^35^, anxiety (State-Trait Anxiety Inventory [STAI])^36^, autonomic dysfunction (Scales for Outcomes in Parkinson’s Disease-Autonomic [SCOPA-AUT])^37^, olfactory function (University of Pennsylvania Smell Identification Test [UPSIT])^38^, daytime sleepiness (Epworth Sleepiness Scale [ESS])^39^, REM sleep behavior disorder symptoms (REM Sleep Behavior Disorder Screening Questionnaire [RBDSQ])^40^, and functional status (Schwab and England Activities of Daily Living Scale). Tremor and PIGD scores were derived from the corresponding MDS-UPDRS Part III items, and apathy was assessed^41^ using the MDS-UPDRS Part I apathy item. Composite cognitive domain scores were calculated by averaging standardized neuropsychological test scores within each domain^42,43^. Executive function was assessed using Letter Number Sequencing and the Symbol Digit Modalities Test; memory was assessed using Hopkins Verbal Learning Test total recall, delayed recall, retention, and recognition discrimination; language was assessed using Semantic Fluency; and visuospatial function was assessed using the Benton Judgment of Line Orientation test^43,44^. A global cognitive composite score was calculated as the mean of the four domain composite scores. Variable definitions and scoring followed standardized PPMI procedures and were harmonized using our open-source data wrangling pipeline.

### 2.3 Classification of Cognitive Status

Participants were classified as converter versus cognitively stable PD based on longitudinal cognitive assessments over a maximum follow-up period of six years. Longitudinal cognitive status was determined using the PPMI Cognitive_State variable, which represents the investigator-assigned cognitive diagnosis based on clinical assessments^30^. Participants were classified as cognitively stable if they remained cognitively normal (Cognitive_state = 1) at all available visits during the follow-up period, and PD-converters if they transitioned from cognitively normal at first visit to MCI or dementia (Cognitive_state = 2 or 3) during follow-up, and remained impaired thereafter without reverting back to normal cognition. Participants who showed fluctuating cognitive status^45^ (i.e. reverted back to normal cognition in future follow-ups) were excluded.

### 2.4 MRI processing

T1-weighted MRIs were processed using PELICAN^46^, an in-house, open-source image processing pipeline. Preprocessing steps included denoising^47^, correction for intensity non-uniformity^48^, and intensity normalization to a standardized range. All images were first linearly and then nonlinearly registered to the MNI-ICBM152 average template^49,50^. Visual quality control was performed for all preprocessing and registration steps by an experienced rater (RM) blinded to clinical outcomes, and scans that failed quality control were excluded from subsequent analyses (N = 69 visits)^51^. Deformation-based morphometry (DBM) was used to quantify regional gray matter atrophy, calculated as the Jacobian determinant of the nonlinear deformation field^52^. DBM values reflect the relative volume of each voxel compared to the MNI-ICBM152 template, where a value below 1 indicates relative volume loss (i.e. atrophy), a value above 1 indicates expansion, and 1 denotes the same volume compared to its equivalent voxel in the template. Average DBM values were extracted from 102 cortical and subcortical regions defined by the CerebrA atlas^53^. WMHs were segmented as part of the PELICAN pipeline using BISON^54^, a previously validated automated tissue segmentation tool capable of identifying WMHs using T1-weighted images. WMH segmentations were visually quality controlled, and scans failing quality control were excluded (N = 56 visits)^54^. Global and lobar WMH volumes were calculated in the stereotaxic space using lobar and hemispheric parcellations (based on Hammer’s lobar atlas)^54,55^. WMH volumes were log-transformed to normalize their distribution prior to statistical analyses.

### 2.5 Statistical analyses

Baseline demographic and clinical characteristics were compared between cognitively stable and converter PD participants using independent-samples t-tests for continuous variables and chi-square tests for categorical variables. Linear mixed-effects models were used to examine potential differences in longitudinal trajectories of DBM (voxel-wise and regional) and global and regional WMH measures between PD converters and cognitively stable PD participants.

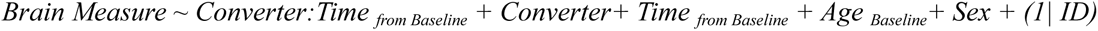

Where *Brain Measure* refers to DBM or WMH measures, *Converter* is a categorical variable indicating participants’ conversion status, and *Time _from Baseline_* indicates the time between the baseline visit and each MRI assessment. The primary variable of interest was *Converter:Time _from Baseline_* interaction, which captures the differences in the rates of change between converter and stable participants (i.e. changes observed in the converters that are above and beyond what is also present in stable PD participants). In addition to interaction effects, the main effects of conversion status and *Time _from Baseline_* were examined, where conversion status reflects the structural differences between PD converters and cognitively stable PD participants already present at baseline and *Time _from Baseline_* reflects the longitudinal changes in stable PD participants. Participant ID was included as a random intercept to account for repeated measurements within participants. All analyses were corrected for multiple comparisons using the False Discovery Rate (FDR) controlling method^56^.

Similar longitudinal linear mixed-effects models were applied to clinical measures, including executive function, memory, language, visuospatial function, and the global cognitive composite score. Additional models were further adjusted for baseline MoCA to ensure findings were not primarily driven by differences in baseline global cognitive performance. As an additional sensitivity analysis, longitudinal clinical models were further adjusted for levodopa equivalent daily dose (LEDD) to assess the potential influence of dopaminergic medication exposure on the observed *Converter:Time _from Baseline_*effects.

To derive an individual-level measure of the spatial structural pattern associated with cognitive conversion in PD, a PD conversion-related DBM pattern expression score was calculated using the voxelwise conversion-status effect map. For each participant, DBM values were multiplied by the corresponding voxelwise t-statistic weights and averaged to obtain a single DBM pattern expression score. Baseline DBM pattern expression score was calculated from each participant’s baseline MRI and therefore remained constant across longitudinal clinical observations. Baseline total WMH burden was similarly obtained from the baseline MRI. Log-transformed total WMH burden and DBM pattern expression score were used in subsequent analyses, examining the associations between the two baseline imaging measures and longitudinal progression in different clinical outcomes using linear mixed-effects models:

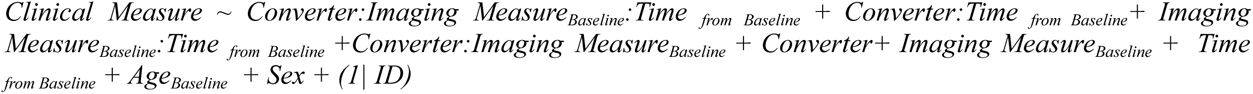

where the Imaging Measure represented either baseline log-transformed total WMH burden or baseline log-transformed DBM pattern expression score. The primary term of interest was the *Converter:Imaging Measure_Baseline_:Time _from Baseline_*, reflecting whether the association between the baseline imaging measure and progression of longitudinal clinical symptoms differed between future PD converters and cognitively stable participants.

To determine whether baseline WMH burden and DBM pattern expression score showed statistically independent associations with subsequent clinical progression, both imaging measures were subsequently included within the same mixed-effects model:

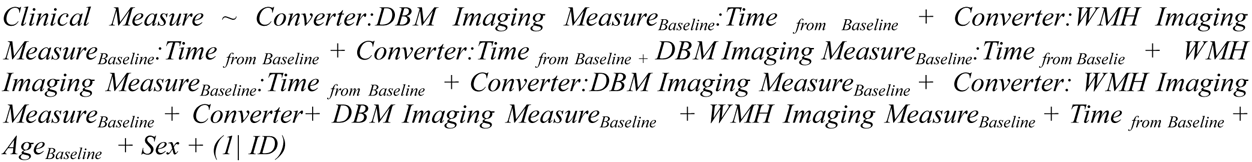

The primary terms of interest were the *Converter:DBM Imaging Measure_Baseline_:Time _from Baseline_ and Converter:WMH Imaging Measure_Baseline_:Time _from Baseline_* interactions. Because both imaging measures were included simultaneously, each coefficient estimated its association with differential longitudinal clinical change while accounting for the other imaging measure. For visualization of significant three-way interactions, participants were divided into lower and higher Imaging Measure groups using the median of the corresponding baseline Imaging Measure separately within each conversion-status group. Predicted trajectories were evaluated at the median value of each subgroup and presented with pointwise 95% confidence intervals. Median splitting was used for visualization only; all statistical inference was based on the continuous imaging measures.

To reduce the potential effect of baseline age differences between groups (as PD converters were significantly older than cognitively stable PD participants at baseline), the groups were age matched based on baseline information. All converter participants were retained, and a subset of stable participants was selected to create groups that are comparable in terms of age. Sex was not used as a strict matching criterion but was evaluated after matching to confirm that the sex distribution remained comparable between groups. Supplementary Figure 2 shows the age distributions before and after matching. As a sensitivity analysis, clinical measure models were subsequently re-estimated in the age-matched imaging sample using the same model specification, covariates, standardization procedure, and FDR correction as in the primary analyses. Baseline age remained included as a covariate in these models to account for residual age variability after matching.

## 3. Results

### 3.1 Participant characteristics

Demographic and clinical characteristics of the study sample are summarized in Table 1. Over the examined follow-up period, 58 participants experienced cognitive decline and remained cognitively impaired thereafter, while 292 participants remained cognitively stable. Participants with fluctuating or inconsistent cognitive status (N = 61) were excluded (Figure S1). Prior to matching, PD converters were older than cognitively stable participants (67.20 ± 6.16 vs. 59.82 ± 9.73 years, p <0.001). PD converters also demonstrated higher MDS-UPDRS Part III, tremor, PIGD, apathy, GDS, SCOPA-AUT, STAI, and RBDSQ scores compared with cognitively stable PD participants. After age-matching, age differences between groups were no longer significant (67.20 ± 6.16 vs. 66.12 ± 5.90 years, p = 0.242) (Figure S2). Differences in PIGD, apathy, GDS, SCOPA-AUT, STAI, and RBDSQ scores remained significant after age matching, whereas baseline differences in MDS-UPDRS Part III and tremor scores were no longer significant.

**Table 1.** Demographic and clinical characteristics of cognitively stable participants and PD converters before and after nearest-neighbor age matching at baseline.

| Measure | PD Cognitively stable | PD converters | P | PD Cognitively stable | PD converters | P |
| --- | --- | --- | --- | --- | --- | --- |
| Stage | **Before matching** |  |  | **After matching** |  |  |
| N | **292 vs 58** |  |  | **174 vs 58** |  |  |
| Age (years) | 59.82 ± 9.73 | 67.20 ± 6.16 | <0.001 | 66.12 ± 5.90 | 67.20 ± 6.16 | 0.242 |
| Female sex (%) | 36.60% | 32.80% | 0.573 | 39.10% | 32.80% | 0.389 |
| Education (years) | 16.09 ± 2.95 | 15.84 ± 2.65 | 0.537 | 15.85 ± 2.97 | 15.84 ± 2.65 | 0.981 |
| MoCA | 27.37 ± 2.12 | 26.72 ± 2.33 | 0.053 | 26.85 ± 2.22 | 26.72 ± 2.33 | 0.718 |
| MDS-UPDRS Part III | 19.90 ± 8.71 | 23.28 ± 10.68 | 0.027 | 21.14 ± 9.26 | 23.28 ± 10.68 | 0.177 |
| Tremor score | 0.73 ± 0.39 | 0.91 ± 0.53 | 0.019 | 0.76 ± 0.36 | 0.91 ± 0.53 | 0.052 |
| PIGD score | 0.19 ± 0.20 | 0.29 ± 0.36 | 0.047 | 0.19 ± 0.20 | 0.29 ± 0.36 | 0.038 |
| Apathy score | 0.16 ± 0.48 | 0.43 ± 0.77 | 0.012 | 0.16 ± 0.50 | 0.43 ± 0.77 | 0.013 |
| GDS score | 1.76 ± 2.19 | 3.05 ± 3.16 | 0.004 | 1.61 ± 2.01 | 3.05 ± 3.16 | 0.002 |
| SCOPA-AUT | 8.22 ± 5.04 | 14.04 ± 8.45 | <0.001 | 9.09 ± 5.10 | 14.04 ± 8.45 | <0.001 |
| UPSIT | 14.31 ± 17.49 | 18.05 ± 19.17 | 0.176 | 16.14 ± 17.99 | 18.05 ± 19.17 | 0.511 |
| STAI total | 60.99 ± 17.18 | 68.67 ± 20.39 | 0.01 | 58.60 ± 16.09 | 68.67 ± 20.39 | 0.001 |
| H&Y stage | 1.57 ± 0.50 | 1.54 ± 0.50 | 0.661 | 1.65 ± 0.49 | 1.54 ± 0.50 | 0.155 |
| Epworth | 5.03 ± 3.10 | 5.98 ± 3.75 | 0.073 | 5.01 ± 2.97 | 5.98 ± 3.75 | 0.075 |
| RBDSQ | 3.00 ± 2.62 | 4.43 ± 2.72 | <0.001 | 3.01 ± 2.80 | 4.43 ± 2.72 | <0.001 |
| Schwab & England | 94.50 ± 5.50 | 93.36 ± 5.41 | 0.147 | 94.28 ± 5.66 | 93.36 ± 5.41 | 0.271 |

MoCA: Montreal Cognitive Assessment score; MDS-UPDRS Part III: Movement Disorder Society Unified Parkinson's Disease Rating Scale Part III; Tremor score: Tremor subscore derived from the MDS-UPDRS Part III; PIGD: Postural Instability and Gait Difficulty score; Apathy: MDS-UPDRS Part I apathy item; GDS: Geriatric Depression Scale score; SCOPA-AUT: Scales for Outcomes in Parkinson's Disease-Autonomic; UPSIT: University of Pennsylvania Smell Identification Test; STAI total: State-Trait Anxiety Inventory total; H&Y stage: Hoehn and Yahr disease stage; Epworth: Epworth Sleepiness Scale; RBDSQ: REM Sleep Behavior Disorder Screening Questionnaire; Schwab & England: Schwab and England Activities of Daily Living (Independence) Scale. Values are presented as mean ± standard deviation unless otherwise indicated. Group comparisons were performed using independent-samples t-tests for continuous variables and chi-square tests for categorical variables. Bold values indicate $p < 0.05$ .

### 3.2 Longitudinal clinical trajectories

Longitudinal models revealed significant Converter:Time _from Baseline_ interactions across all cognitive domains and MoCA, indicating faster cognitive decline in PD converters compared with cognitively stable PD participants (Table 2). In addition to cognitive decline, PD converters demonstrated faster worsening of MDS-UPDRS Part III, PIGD, Hoehn and Yahr stage, depressive symptoms (GDS), anxiety (STAI), daytime sleepiness (Epworth Sleepiness Scale), and functional status (Schwab and England score). No significant Converter:Time _from Baseline_ interactions were observed for tremor, apathy, REM sleep behavior disorder symptoms (RBDSQ), or autonomic dysfunction (SCOPA-AUT). These findings suggest that cognitive conversion in PD is associated with broader multisystem clinical progression extending beyond cognition alone. Adjustment for baseline MoCA produced minimal changes in the *Converter:Time _from Baseline,_*interaction, whereas additional adjustment for LEDD also produced minimal changes in the overall pattern and magnitude of the effects, indicating that the observed longitudinal differences were largely robust to adjustment for baseline global cognitive performance, and dopaminergic medication exposure (Table S1). The overall pattern of longitudinal group differences was also largely preserved, although the language and MDS-UPDRS Part III interaction were no longer significant, whereas apathy became significant after age matching (Table 2).

**Table 2.** Longitudinal clinical trajectories comparing converter and stable PD participants.

| Measure | $\beta$ | T statistics | P-value | q-FDR | $\beta$ | T statistics | P-value | q-FDR |
| --- | --- | --- | --- | --- | --- | --- | --- | --- |
| Stage | Before matching |  |  |  | After matching |  |  |  |
| Executive | -0.073 | -6.2 | <0.001 | <0.001 | -0.052 | -4.22 | <0.001 | <0.001 |
| Memory | -0.055 | -3.93 | <0.001 | <0.001 | -0.059 | -4.04 | <0.001 | <0.001 |
| Language | -0.037 | -2.7 | 0.007 | 0.01 | -0.026 | -1.7 | 0.089 | 0.108 |
| Visuospatial | -0.05 | -3.66 | <0.001 | <0.001 | -0.045 | -3.23 | 0.001 | 0.002 |
| Global cognition | -0.082 | -6.75 | <0.001 | <0.001 | -0.07 | -5.42 | <0.001 | <0.001 |
| MoCA | -0.088 | -6.89 | <0.001 | <0.001 | -0.069 | -4.95 | <0.001 | <0.001 |
| UPDRS-III | 0.033 | 2.74 | 0.006 | 0.009 | 0.023 | 1.85 | 0.064 | 0.083 |
| Tremor | -0.012 | -1.01 | 0.312 | 0.322 | 0.002 | 0.12 | 0.905 | 0.905 |
| PIGD | 0.112 | 8.83 | <0.001 | <0.001 | 0.088 | 6.89 | <0.001 | <0.001 |
| Hoehn & Yahr | 0.075 | 6.36 | <0.001 | <0.001 | 0.079 | 6.23 | <0.001 | <0.001 |
| Apathy | 0.014 | 1.21 | 0.225 | 0.254 | 0.054 | 4.71 | <0.001 | <0.001 |
| GDS | 0.041 | 3.49 | <0.001 | <0.001 | 0.037 | 2.84 | 0.005 | 0.007 |
| STAI | 0.053 | 4.72 | <0.001 | <0.001 | 0.047 | 3.95 | <0.001 | <0.001 |
| Epworth | 0.028 | 2.6 | 0.009 | 0.012 | 0.029 | 2.53 | 0.011 | 0.016 |
| RBD | -0.01 | -0.99 | 0.322 | 0.322 | 0.002 | 0.19 | 0.845 | 0.898 |
| SCOPA-AUT | -0.012 | -1.27 | 0.206 | 0.25 | -0.007 | -0.75 | 0.451 | 0.511 |
| Schwab & England | -0.139 | -15.38 | <0.001 | <0.001 | -0.098 | -10.54 | <0.001 | <0.001 |

Results from linear mixed-effects models examining the interaction between conversion status and time (*Converter:Time<sub>from Baseline</sub>*). N indicates the number of longitudinal observations included in each model. Positive $\beta$ coefficients indicate a more positive longitudinal slope in PD converters relative to cognitively stable PD participants, whereas negative $\beta$ coefficients indicate a more negative longitudinal slope. The clinical interpretation of coefficient direction depends on the scoring direction of each measure. Models were adjusted for age at baseline and sex, with participant ID included as a random intercept to account for repeated measurements. P-values were corrected for multiple comparisons using the Benjamini-Hochberg false discovery rate (FDR) procedure.

### 3.3 Longitudinal atrophy trajectories

No significant baseline group differences were observed in regional DBM measures. Widespread longitudinal structural changes were observed in both groups, involving extensive cortical, subcortical, cerebellar, and ventricular regions (Figure 1a). Furthermore, we observed significant *Converter:Time _from Baseline_* interaction effects, indicating accelerated gray matter atrophy in PD converters compared with cognitively stable PD participants (Figure 1b). At an uncorrected threshold (p < 0.05), interaction effects were observed in the bilateral accumbens area, amygdala, basal forebrain, and thalamus, right third ventricle, cerebellar white matter, lateral ventricle, as well as the left inferior lateral ventricle, entorhinal cortex, and caudate. After FDR correction, significant interaction effects remained in the bilateral amygdala, thalamus, left accumbens area, inferior lateral ventricle, and entorhinal cortex (Table 3). Models additionally adjusted for baseline MoCA showed similar results, with the significant Converter:Time _from Baseline_ effects remaining after FDR correction (Table S2).

**Figure 1.**
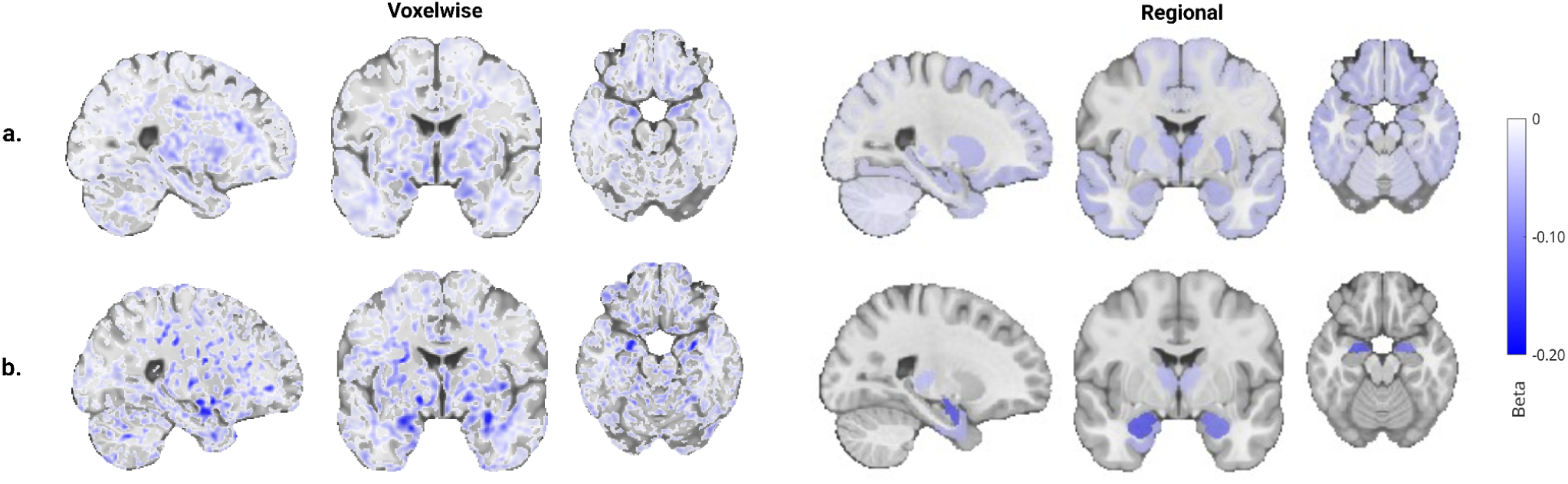
Longitudinal differences in DBM trajectories between PD converters and cognitively stable participants. Presented maps display voxelwise (left) and regional (right) standardized coefficients (β) estimated from linear mixed-effects models. (a) Main effect of Time _from Baseline_ from the model *DBM ∼ Converter:Time _from Baseline_ + Converter+ Time _from Baseline_ + Age _Baseline_+ Sex + (1| ID)*, showing longitudinal gray matter atrophy and ventricular expansion. (b) *Converter:Time _from Baseline_* interaction from the same model, showing regions with accelerated longitudinal gray matter atrophy in PD converters relative to cognitively stable PD participants.

**Table 3.**
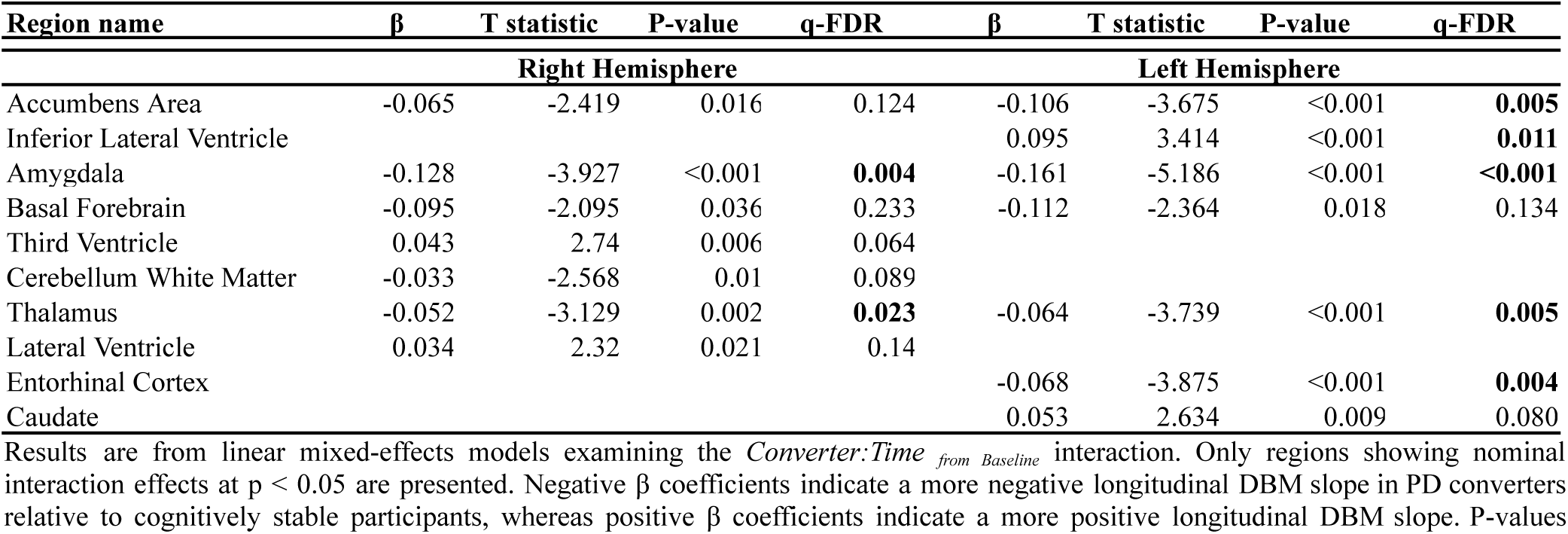
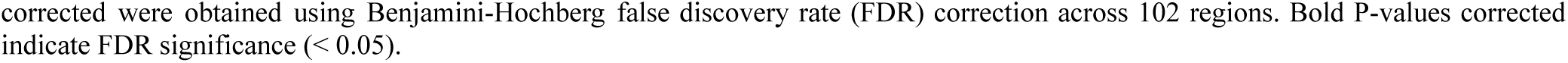
Longitudinal differences in DBM trajectories between PD converters and cognitively stable participants.

| Region name | $\beta$ | T statistic | P-value | q-FDR | $\beta$ | T statistic | P-value | q-FDR |
| --- | --- | --- | --- | --- | --- | --- | --- | --- |
| Right Hemisphere |  |  |  |  | Left Hemisphere |  |  |  |
| Accumbens Area | -0.065 | -2.419 | 0.016 | 0.124 | -0.106 | -3.675 | <0.001 | **0.005** |
| Inferior Lateral Ventricle |  |  |  |  | 0.095 | 3.414 | <0.001 | **0.011** |
| Amygdala | -0.128 | -3.927 | <0.001 | **0.004** | -0.161 | -5.186 | <0.001 | <**0.001** |
| Basal Forebrain | -0.095 | -2.095 | 0.036 | 0.233 | -0.112 | -2.364 | 0.018 | 0.134 |
| Third Ventricle | 0.043 | 2.74 | 0.006 | 0.064 |  |  |  |  |
| Cerebellum White Matter | -0.033 | -2.568 | 0.01 | 0.089 |  |  |  |  |
| Thalamus | -0.052 | -3.129 | 0.002 | **0.023** | -0.064 | -3.739 | <0.001 | **0.005** |
| Lateral Ventricle | 0.034 | 2.32 | 0.021 | 0.14 |  |  |  |  |
| Entorhinal Cortex |  |  |  |  | -0.068 | -3.875 | <0.001 | **0.004** |
| Caudate |  |  |  |  | 0.053 | 2.634 | 0.009 | 0.080 |

Results are from linear mixed-effects models examining the *Converter:Time<sub>from Baseline</sub>* interaction. Only regions showing nominal interaction effects at $p < 0.05$ are presented. Negative $\beta$ coefficients indicate a more negative longitudinal DBM slope in PD converters relative to cognitively stable participants, whereas positive $\beta$ coefficients indicate a more positive longitudinal DBM slope. P-values
corrected were obtained using Benjamini-Hochberg false discovery rate (FDR) correction across 102 regions. Bold P-values corrected indicate FDR significance ( $< 0.05$ ).

The results remained similar after age matching, indicating that the observed atrophy trajectories were not solely attributable to baseline age differences. The effects were localized to the bilateral amygdala and thalamus, left accumbens area, entorhinal cortex, and putamen, as well as the right caudal anterior cingulate cortices (uncorrected P <0.05). After FDR correction, significant interaction effects remained in the bilateral amygdala (Figure S3; Table S3).

### 3.4 Longitudinal WMH trajectories

WMH burden increased significantly over time globally and across frontal, parietal, temporal, and occipital regions in both groups. Linear mixed-effects models revealed no significant Converter:Time _from Baseline_ interactions, but significant main effects of conversion status and time were observed (Figure 2; Table 4). PD converters exhibited greater global and bilateral frontal and left parietal WMH burden than cognitively stable PD participants after FDR correction.

**Figure 2.**
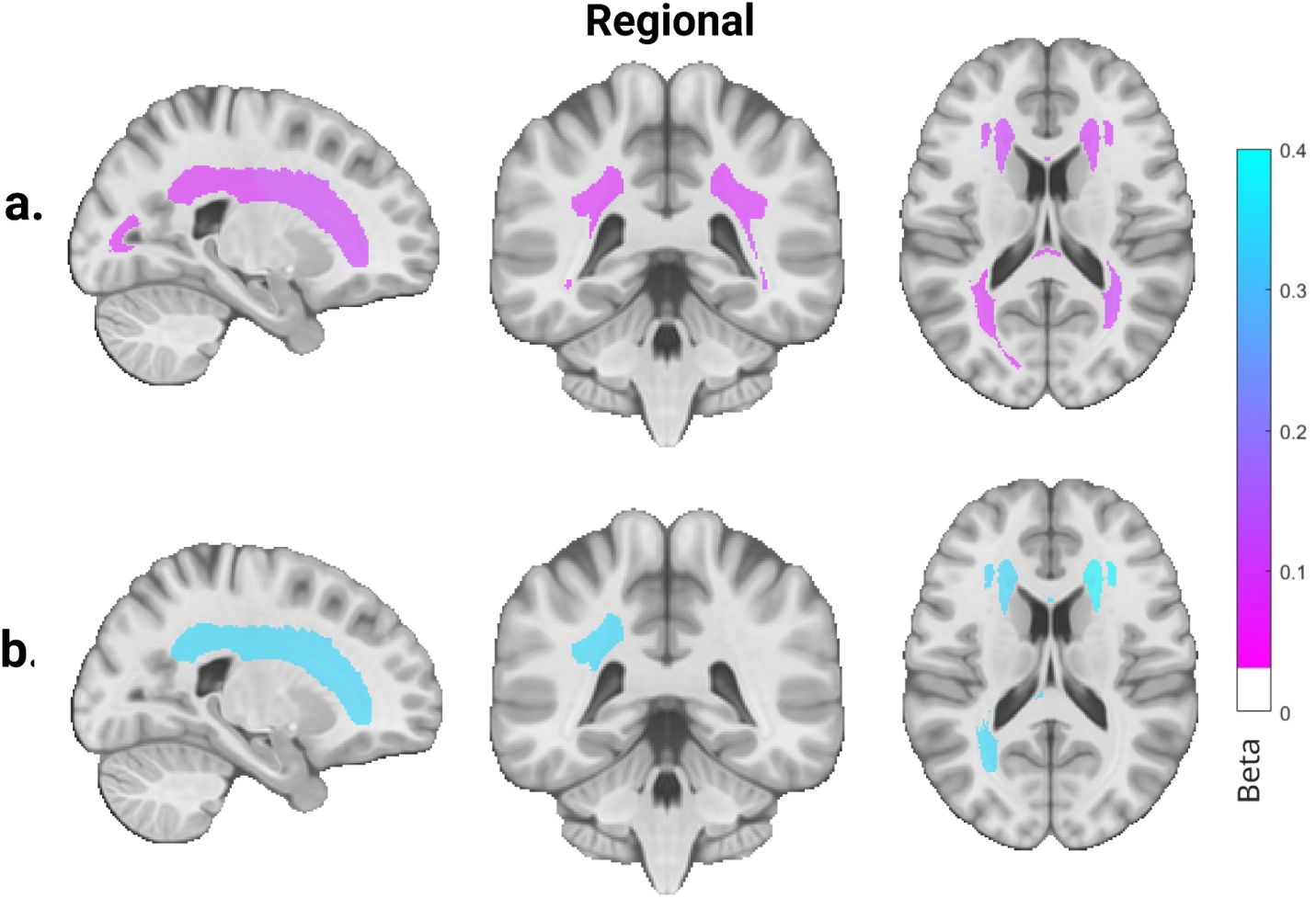
Longitudinal differences in WMH trajectories between PD converters and cognitively stable participants. Presented maps display regional coefficients (β) estimated from linear mixed-effects models. (a) *Time _from Baseline_* estimates from the model *WMH ∼ Converter:Time _from Baseline_ + Converter+ Time _from Baseline_ + Age _Baseline_+ Sex + (1| ID)*, showing longitudinal white matter hyperintensity increase. (b) *Converter* main effects from the same model, showing baseline WMH burden differences in PD converters relative to cognitively stable PD participants.

**Figure 3.**
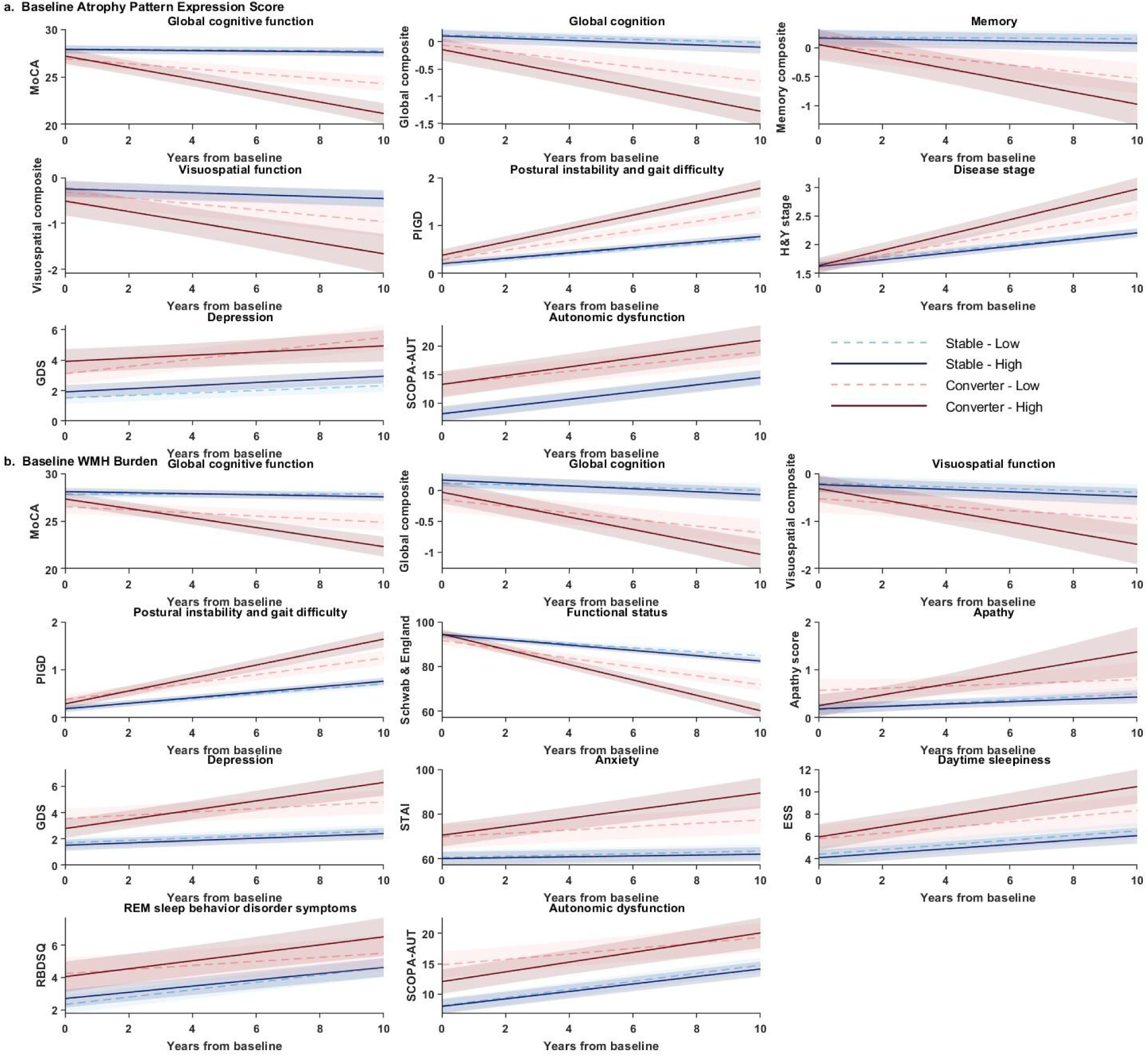
Associations between baseline imaging measures and longitudinal clinical trajectories according to PD conversion status. Predicted longitudinal clinical trajectories are shown according to (a) baseline PD conversion-related atrophy pattern expression score and (b) baseline total white matter hyperintensity (WMH) burden. Predicted clinical trajectories were derived from linear mixed effect models including the *Converter:Imaging Measure_Baseline_:Time _from Baseline_* interaction, adjusted for age and sex, with participant-specific random intercepts. For visualization, low and high imaging groups were defined separately within each conversion-status group using the median of the corresponding baseline imaging measure, and trajectories were evaluated at the median imaging value of each subgroup. For the atrophy analyses, dashed and solid lines indicate low and high atrophy pattern expression scores, respectively; for the WMH analyses, dashed and solid lines indicate low and high WMH burden, respectively. Blue lines represent cognitively stable participants and red lines represent future PD converters. Shaded regions indicate pointwise 95% confidence intervals. Median-based groups were used for visualization only; statistical inference was based on the continuous three-way interaction.

**Table 4.**
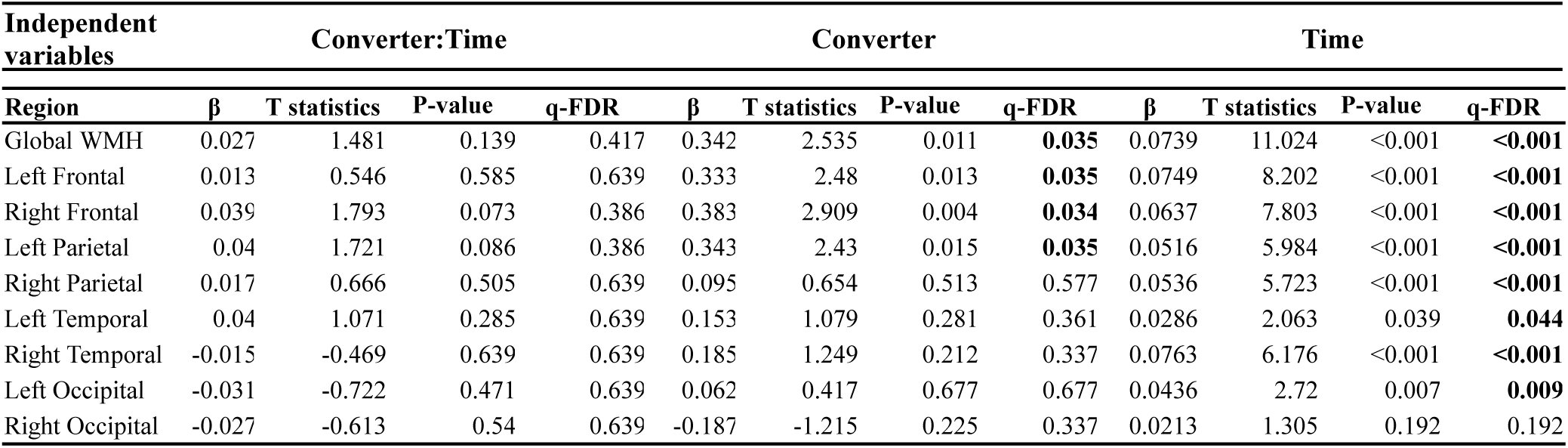
Mixed effects model results comparing longitudinal WMH trajectories between converter and cognitively stable PD participants.

### 3.5 Baseline atrophy pattern and clinical progression

Baseline atrophy pattern expression score showed significant *Converter:DBM_Baseline_:Time _from Baseline_* interactions across several clinical domains, suggesting that a greater expression of the observed baseline atrophy pattern was linked to accelerated subsequent progression in cognitive, gait, disease stage, depressive, and autonomic symptoms (Table S4). Eight outcomes remained significant after FDR correction, including MoCA (β = -0.446, q < 0.001), PIGD (β = 0.236, q < 0.001), Hoehn and Yahr stage (β = 0.237, q < 0.001), global cognition (β = -0.227, q < 0.001), GDS (β = -0.242, q < 0.001), visuospatial function (β = -0.182, q = 0.021), memory (β = -0.170, q = 0.038), and SCOPA-AUT (β = 0.119, q = 0.038). The largest standardized interaction effect was observed for MoCA, indicating a particularly strong difference in the association between baseline DBM pattern expression and subsequent MoCA change between conversion groups. Similarly, for all other domains, the three-way interaction indicated that the converter-stable difference in longitudinal symptom trajectories increased with greater baseline DBM pattern expression, with predicted trajectories showing steeper increases among high-expression converters than low-expression converters.

### 3.6 Baseline WMH burden and clinical progression

Baseline WMH burden was associated with subsequent progression across cognitive, gait, neuropsychiatric, autonomic, sleep-related, and functional domains, with these associations differing according to domain and future conversion status (Table S4). Significant *Converter:WMH_Baseline_:Time _from Baseline_* interactions survived FDR correction for 11 clinical outcomes. Greater baseline WMH burden was associated with more unfavorable longitudinal trajectories in future PD converters relative to cognitively stable participants for MoCA (β = -0.237, q < 0.001), global cognition (β = -0.117, q = 0.046), visuospatial function (β = -0.156, q = 0.024), PIGD (β = 0.179, q < 0.001), GDS (β = 0.228, q < 0.001), SCOPA-AUT (β = 0.151, q = 0.005), Epworth Sleepiness Scale (β = 0.133, q = 0.030), Schwab and England functional status (β = -0.255, q < 0.001), STAI (β = 0.180, q = 0.005), RBDSQ (β = 0.135, q = 0.024), and apathy (β = 0.150, q = 0.030). The positive three-way interactions indicated that the converter-stable difference in longitudinal slope became more evident with increasing baseline WMH burden. No FDR-significant three-way interactions were observed for Hoehn and Yahr stage, memory, language, executive function, tremor, MDS-UPDRS Part III, or olfactory function.

### Sensitivity Analyses within Age-matched Subsamples

Age-matched sensitivity analyses generally supported the primary three-way interaction findings, although several associations were attenuated in the matched sample (Table S5). For baseline WMH burden, significant *Converter:WMH_Baseline_:Time _from Baseline_* interactions remained for MoCA, PIGD, apathy, depressive symptoms, anxiety, autonomic dysfunction, daytime sleepiness, REM sleep behavior disorder symptoms, and Schwab and England functional status. Associations with global cognition and visuospatial function no longer survived FDR correction after age matching. For baseline DBM pattern expression score, significant *Converter:DBM_Baseline_:Time _from Baseline_* interactions remained for MoCA, global cognition, PIGD, Hoehn and Yahr stage, and depressive symptoms. Associations with memory, visuospatial function, and autonomic dysfunction were attenuated and no longer survived FDR correction.

### 3.7 Joint WMH and atrophy models

Baseline WMH burden and atrophy pattern expression scores were moderately correlated (r = 0.40, p = <0.001). To determine whether the imaging measures retained associations with longitudinal clinical progression after accounting for the other measure, they were simultaneously included in the models. FDR-significant *Converter:WMH_Baseline_:Time _from Baseline_* interactions remained for GDS (β = 0.395, q < 0.001), STAI (β = 0.241, q < 0.001), and Schwab and England functional status (β = -0.246, q < 0.001), and significant *Converter:DBM_Baseline_:Time _from Baseline_* interactions remained for MoCA (β = -0.395, q < 0.001), PIGD (β = 0.179, q = 0.005), Hoehn and Yahr stage (β = 0.233, q = 0.002), global cognition (β = -0.195, q = 0.008), GDS (β = -0.419, q < 0.001), memory (β = -0.182, q = 0.047), and STAI (β = -0.177, q = 0.025). Thus, after mutual adjustment, atrophy pattern expression score retained broader associations with cognitive and motor progression, whereas WMH burden retained more selective associations with neuropsychiatric symptoms and functional independence (Table S6).

## 4. Discussion

In this study, we investigated the longitudinal structural brain changes and clinical trajectories associated with conversion from normal to impaired cognition in de novo PD. Future converters exhibited accelerated longitudinal gray matter atrophy, with prominent involvement of limbic and subcortical regions, consistent with prior studies^8–10,12^. Baseline WMH burden and atrophy pattern expression score were also associated with subsequent clinical trajectories in a conversion-status-dependent manner. Joint analyses further indicated that these vascular and neurodegenerative imaging measures provide partially overlapping but complementary information about subsequent multidomain clinical progression.

Beyond cognitive decline, future converters showed a broader pattern of clinical progression involving gait and disease stage, depressive and anxiety symptoms, daytime sleepiness, and functional independence. Cognitive decline was evident across executive, memory, language, visuospatial, and global cognitive measures, indicating that conversion was not driven by a single cognitive domain. This broader pattern is consistent with previous longitudinal studies showing that cognitive vulnerability in PD is associated with faster motor, gait, non-motor, and functional progression^25–27^. Depression and anxiety have also been linked to subsequent cognitive decline in PD^28^. The persistence of most of these differences after age matching further supports a multidomain progressive phenotype associated with cognitive conversion.

Limbic and subcortical structures showed accelerated neurodegeneration associated with future cognitive decline status. Future converters exhibited faster atrophy within the bilateral amygdala and thalamus, as well as the left nucleus accumbens and entorhinal cortex, together with greater ventricular expansion. Bilateral amygdala atrophy results remained significant after age matching and FDR correction, indicating that the association was not explained solely by baseline age differences. These findings are consistent with growing evidence suggesting that limbic neurodegeneration is detectable before cognitive impairment becomes clinically apparent in PD^4,6,11^. Accelerated entorhinal atrophy is also consistent with the early involvement of medial temporal structures in PD-related cognitive impairment^4,11^. Similarly, accelerated thalamic atrophy supports growing evidence implicating thalamic and striatal structures in cognitive deterioration in PD^4,6,12^. The persistence of the effects after additional adjustment for baseline MoCA further suggests that these structural differences were not primarily explained by initial global cognitive performance.

Beyond identifying regional structural changes associated with future cognitive conversion, we quantified the expression of a distributed PD conversion-related atrophy pattern at the individual level and examined whether its baseline expression was associated with subsequent clinical progression. Significant interactions were observed across cognitive, motor, and non-motor domains, including MoCA, global cognition, memory, visuospatial function, PIGD, Hoehn and Yahr stage, depressive symptoms, and autonomic dysfunction. These associations suggest that the observed distributed atrophy pattern is associated with broader disease progression rather than cognitive decline alone, consistent with previous longitudinal studies demonstrating that cognitive impairment in PD is accompanied by worsening motor, functional, and non-motor outcomes^25–27^. Since the atrophy pattern expression score summarizes the weighted expression of the voxelwise PD conversion-related atrophy pattern, it provides a subject-level measure of the distributed structural abnormalities rather than isolated regional abnormalities.

Although future PD converters exhibited greater overall WMH burden than cognitively stable participants, they did not demonstrate significantly faster longitudinal WMH accumulation. This contrasts with our gray matter findings, in which future PD converters showed accelerated longitudinal atrophy but not baseline differences, suggesting that gray matter neurodegeneration and cerebrovascular pathology may have distinct temporal contributions to cognitive decline. Importantly, baseline WMH burden was associated with subsequent clinical trajectories across a broad range of domains, including cognition, gait impairment, neuropsychiatric symptoms, autonomic dysfunction, sleep-related symptoms, and functional independence. For example, the significant *Converter:Imaging Measure_Baseline_:Time _from Baseline_* interaction indicated that the association between baseline WMH burden and longitudinal autonomic change differed by conversion status, with predicted trajectories showing steeper SCOPA-AUT increases among converters with greater baseline WMH burden, and this association remained significant after age matching. Our findings suggest that WMHs may represent pre-existing cerebrovascular vulnerability that modifies subsequent clinical progression rather than a process that shows accelerated accumulation in future PD converters. This interpretation is consistent with longitudinal evidence showing that baseline WMH burden predicts subsequent cognitive decline and progression of cognitive impairment in de novo and early PD^17,18^, whereas early cross-sectional studies found no significant association between WMH burden and cognitive function in newly diagnosed untreated PD^16^, and longitudinal WMH accumulation has been reported to show limited association with cognitive change in PD^21^. Together, these findings suggest that WMHs may reflect an underlying vulnerability to cognitive decline^13,17^, whereas accelerated gray matter atrophy may more closely reflect the progressive structural neurodegeneration associated with future cognitive conversion in PD^8–10^.

Joint atrophy and WMH models further supported the presence of overlapping and complementary information provided by baseline WMH burden and atrophy pattern expression score. When both imaging measures were considered simultaneously, DBM pattern expression score retained significant associations with MoCA, global cognition, memory, PIGD, Hoehn and Yahr stage, depressive symptoms, and anxiety, supporting broader associations across cognitive, motor, and neuropsychiatric domains. WMH burden retained associations with depressive symptoms, anxiety, and functional independence. These findings are consistent with previous evidence linking cerebrovascular burden to neuropsychiatric symptoms in PD^57^.

Taken together, the temporal clinical patterns observed in this study support a model in which cerebrovascular and neurodegenerative abnormalities may relate differently to cognitive vulnerability in PD. WMH burden was already greater in future converters but did not subsequently accumulate at a faster rate, consistent with a background vulnerability that may lower resilience to progressive neurodegeneration^20,24^. In contrast, future converters demonstrated accelerated longitudinal gray matter atrophy, while baseline expression of the distributed atrophy pattern retained particularly broad associations with subsequent cognitive and motor progression. The joint models further suggest that these imaging measures are neither completely independent nor redundant, with some clinical associations shared and others preferentially retained by one imaging measure after mutual adjustment. Together, these cerebrovascular and neurodegenerative abnormalities may contribute to the heterogeneity of subsequent cognitive and multisystem clinical trajectories in early PD.

Our study has several important strengths. We investigated a large longitudinal cohort of newly diagnosed, untreated PD participants who were cognitively normal at baseline, enabling characterization of structural changes associated with subsequent cognitive conversion before the confounding effects of dopaminergic treatment or advanced disease progression. By integrating longitudinal DBM analyses, WMH quantification, and comprehensive clinical assessments within the same cohort, we characterized both gray matter neurodegeneration and cerebrovascular pathology associated with future cognitive conversion in PD. Furthermore, age-matched sensitivity analyses and additional adjustment for baseline MoCA further supported the robustness of the observed findings.

From a clinical perspective, given the established impact of cognitive impairment on disability and quality of life^58,59^, these findings suggest risk stratification for cognitive decline in early PD may benefit from considering both neurodegenerative and cerebrovascular pathways. The absence of accelerated WMH accumulation should not be interpreted as evidence that vascular pathology is clinically unimportant; rather, baseline vascular burden may identify a background vulnerability upon which progressive neurodegeneration acts. Conversely, longitudinal gray matter change may provide information about the active neurodegenerative processes more proximally associated with cognitive conversion.

Certain limitations should be considered when interpreting our findings. First, although our longitudinal design provides stronger evidence for temporal associations than cross-sectional studies, the observational nature of the study precludes conclusions regarding the causal mechanisms linking regional atrophy to cognitive decline. Second, despite studying a large longitudinal cohort of de novo PD participants, the number of individuals who experienced cognitive decline remained substantially smaller than the cognitively stable group, which may have limited our ability to detect more subtle longitudinal regional effects after correction for multiple comparisons. Third, MCI and dementia were combined into a single cognitive-decline group to maximize the converter sample size and because dementia classifications were often not stable or occurred beyond the 6-year analysis window. For example, among participants diagnosed with dementia at any visit (N = 25), 48% received the diagnosis after 6 years, 36% were subsequently reclassified as MCI, and 16% received the diagnosis only at their final visit, precluding assessment of diagnostic stability. Future studies with longer follow-ups are therefore needed to distinguish MCI and dementia trajectories. Replication in large, independent longitudinal cohorts will also be important to confirm the robustness and generalizability of these findings.

In summary, this study demonstrates that accelerated longitudinal gray matter atrophy is associated with subsequent cognitive conversion in PD. Bilateral amygdala atrophy emerged as the most robust age-independent structural imaging marker of future cognitive conversion within our cohort. Baseline expression of the distributed PD conversion-related atrophy pattern was associated with subsequent cognitive, motor, and non-motor trajectories, indicating that structural vulnerability extends beyond isolated cognitive decline. In contrast, although future converters exhibited greater baseline WMH burden, they did not demonstrate accelerated longitudinal WMH accumulation, suggesting that cerebrovascular pathology and gray matter neurodegeneration may have distinct temporal patterns in relation to cognitive decline. Importantly, baseline WMH burden and DBM pattern expression score were also associated with subsequent multidomain clinical trajectories, and joint models indicated overlapping but complementary associations after mutual adjustment. DBM pattern expression score retained broader associations with cognitive and motor progression, whereas WMH burden retained associations with neuropsychiatric symptoms and functional independence. Together, these findings highlight the value of longitudinal neuroimaging for improving our understanding of early cognitive conversion in PD and provide further insight into the neurobiological mechanisms underlying cognitive decline.

## Acknowledgments

Data used in this article were obtained from the Parkinson’s Progression Markers Initiative (PPMI) database ([www.ppmi-info.org/data](http://www.ppmi-info.org/data)). For up-to-date information on the study, visit [www.ppmi-info.org](http://www.ppmi-info.org). PPMI is sponsored and partially funded by The Michael J. Fox Foundation for Parkinson’s Research and funding partners, including AbbVie, Avid Radiopharmaceuticals, Biogen, Bristol-Myers Squibb, Covance, GE Healthcare, Genentech, GlaxoSmithKline (GSK), Eli Lilly and Company, Lundbeck, Merck, Meso Scale Discovery (MSD), Pfizer, Piramal Imaging, Roche, Servier, and UCB ([www.ppmi-info.org/fundingpartners](http://www.ppmi-info.org/fundingpartners)). The authors also acknowledge the use of Digital Research Alliance of Canada (alliancecan.ca) resources for performing the image-processing analyses presented in this work.

## Data availability

Data used in the preparation of this manuscript were obtained from the Parkinson’s Progression Markers Initiative (PPMI) database (https://www.ppmi-info.org/access-data-specimens/download-data). All the information and data used in this study are publicly available for free and can be requested on the PPMI website (https://www.ppmi-info.org).

